# Signals of significantly increased vaccine breakthrough, decreased hospitalization rates, and less severe disease in patients with COVID-19 caused by the Omicron variant of SARS-CoV-2 in Houston, Texas

**DOI:** 10.1101/2021.12.30.21268560

**Authors:** Paul A. Christensen, Randall J. Olsen, S. Wesley Long, Richard Snehal, James J. Davis, Matthew Ojeda Saavedra, Kristina Reppond, Madison N. Shyer, Jessica Cambric, Ryan Gadd, Rashi M. Thakur, Akanksha Batajoo, Regan Mangham, Sindy Pena, Trina Trinh, Jacob C. Kinskey, Guy Williams, Robert Olson, Jimmy Gollihar, James M. Musser

## Abstract

Genetic variants of SARS-CoV-2 continue to dramatically alter the landscape of the COVID-19 pandemic. The recently described variant of concern designated Omicron (B.1.1.529) has rapidly spread worldwide and is now responsible for the majority of COVID-19 cases in many countries. Because Omicron was recognized very recently, many knowledge gaps exist about its epidemiology, clinical severity, and disease course. A genome sequencing study of SARS-CoV-2 in the Houston Methodist healthcare system identified 4,468 symptomatic patients with infections caused by Omicron from late November 2021 through January 5, 2022. Omicron very rapidly increased in only three weeks to cause 90% of all new COVID-19 cases, and at the end of the study period caused 98% of new cases. Compared to patients infected with either Alpha or Delta variants in our healthcare system, Omicron patients were significantly younger, had significantly increased vaccine breakthrough rates, and were significantly less likely to be hospitalized. Omicron patients required less intense respiratory support and had a shorter length of hospital stay, consistent with on average decreased disease severity. Two patients with Omicron “stealth” sublineage BA.2 also were identified. The data document the unusually rapid spread and increased occurrence of COVID-19 caused by the Omicron variant in metropolitan Houston, and address the lack of information about disease character among US patients.

## [Introduction]

Over the last 14 months, the Alpha and Delta variants of concern (VOCs) of SARS-CoV-2 have caused two distinct COVID-19 disease surges in the United States, Southeast Asia, Europe, and elsewhere (https://www.cdc.gov/coronavirus/2019-ncov/cases-updates/variant-surveillance/variant-info.html, last accessed December 30, 2021; https://www.gov.uk/government/collections/new-sars-cov-2-variant, last accessed December 30, 2021), and remodeled the landscape of human behavior and many societies. Delta replaced the Alpha variant as the cause of virtually all COVID-19 in many countries (https://www.who.int/publications/m/item/weekly-epidemiologicalupdate-on-covid-19---13-july-2021, last accessed August 18, 2021; https://www.ons.gov.uk/peoplepopulationandcommunity/healthandsocialcare/conditionsanddiseases/bulletins/coronaviruscovid19infectionsurveypilot/9july2021, last accessed August 18, 2021).

At the start of the pandemic almost two years ago, the Houston Methodist healthcare system instituted a comprehensive and integrated population genomics project designed to sequence all SARS-CoV-2 samples causing COVID-19 in patients cared for at our facilities, which include eight hospitals located throughout the metroplex. The project was implemented when the initial Houston Methodist COVID-19 case was diagnosed at the end of February 2020, and has continued unabated^1-7^. This project was facilitated by the existence of a single large diagnostic laboratory that serves the entire system and is seamlessly integrated with a research institute with extensive genomics expertise and capacity. A key goal was to comprehensively map the population genomics, trajectory, and other features of the pandemic in metropolitan Houston with a population size of approximately 7.2 million. Houston is the fourth largest city in the United States, the most ethnically diverse metropolitan area in the country, and is a major port of entry. To date, SARS-CoV-2 genomes have been sequenced from greater than 70,000 patient samples. Many features of four distinct SARS-CoV-2 waves in Houston have been described^2-6^.

The successes of rapid SARS-CoV-2 vaccine development and documented efficacy, coupled with the significant downturn of the disease wave caused by Delta in Houston and elsewhere in fall, 2021^6^, suggested that the pandemic was abating. However, the identification of a new VOC designated B.1.1.529 and known as Omicron that has spread rapidly in South Africa and the UK has tempered this optimism^8-10^. Inasmuch as Omicron was recognized very recently, and much is not known about its epidemiology and clinical characteristics and course, we used our integrated infrastructure in an effort to address the lack of information available for United States Omicron patients. Genome sequencing identified 4,468 COVID-19 patients with symptomatic disease caused by Omicron in the Houston Methodist healthcare system beginning in late November 2021 and ending January 5, 2022. In three weeks Omicron spread throughout the Houston metropolitan region to become the cause of 90% of new COVID-19 cases, and at the end of the study period caused 98% of all new cases. Compared to patients infected with either Alpha or Delta variants and cared for in our system, significantly fewer Omicron patients were hospitalized, and those who were hospitalized required significantly less intense respiratory support and had a shorter length of stay. Our findings are consistent with decreased disease severity among Houston Methodist Omicron patients. Many factors undoubtedly have contributed, including but not limited to increased vaccination uptake, population immunity, and patient demographics such as younger age. The extent to which our findings translate to other cities and other patient populations, including children, is unknown. These data expand on our initial Omicron work^7^ and address the lack of information about disease character among US patients with COVID-19 caused by this VOC.

## Materials and Methods

### Patient Specimens

Specimens were obtained from patients registered at Houston Methodist facilities (e.g., hospitals and urgent care centers), and institutions in the Houston metropolitan region that use our laboratory services. The great majority of individuals had signs or symptoms consistent with COVID-19 disease. For analyses focusing on patients with COVID-19 caused by the Omicron variant, samples obtained from November 27, 2021 through January 5, 2022 were used. This time frame was chosen because it represents the period during which an Omicron variant was first identified in our healthcare system and the last date of specimen collection used to generate genome sequence data for this manuscript. Note that the genome data were generated for two distinct sampling periods. The first period included November 27, 2021 through December 23, 2021 and the second period included samples obtained between December 30, 2021 through January 5, 2022. This discontinuous sampling strategy was used in an effort to obtain the most up-to-date data available for inclusion in this study. Because of the substantial number of positive specimens obtained daily in the December 24, 2021 to December 29, 2021 period (sometimes exceeding 1,500) it wasn’t possible to sequence most of the samples collected during this period for inclusion in the study.

For analyses comparing features of patients infected with the Omicron VOC and Alpha and Delta VOCs, all patients documented to be infected with these variants in the Houston Methodist system were studied. The study included 40,991 unique patients identified in this time frame for whom we had SARS-CoV-2 genome sequences. The work was approved by the Houston Methodist Research Institute Institutional Review Board (IRB1010-0199).

### SARS-CoV-2 Molecular Diagnostic Testing

Specimens obtained from symptomatic patients with a suspicion for COVID-19 disease were tested in the Molecular Diagnostics Laboratory at Houston Methodist Hospital using assays granted Emergency Use Authorization (EUA) from the FDA (https://www.fda.gov/medical-devices/emergency-situations-medical-devices/faqs-diagnostic-testing-sars-cov-2#offeringtests, last accessed June 7, 2021). Multiple molecular testing platforms were used, including the COVID-19 test or RP2.1 test with BioFire Film Array instruments, the Xpert Xpress SARS-CoV-2 test using Cepheid GeneXpert Infinity or Cepheid GeneXpert Xpress IV instruments, the Cobas SARS-CoV-2 & Influenza A/B Assay using the Roche Liat system, the SARS-CoV-2 Assay using the Hologic Panther instrument, the Aptima SARS-CoV-2 Assay using the Hologic Panther Fusion system, the Cobas SARS-CoV-2 test using the Roche 6800 system, and the SARS-CoV-2 assay using Abbott Alinity m instruments. Virtually all tests were performed on material obtained from nasopharyngeal swabs immersed in universal transport media (UTM); oropharyngeal or nasal swabs, bronchoalveolar lavage fluid, or sputum treated with dithiothreitol (DTT) were sometimes used. Standardized specimen collection methods were used (https://vimeo.com/396996468/2228335d56, last accessed June 7, 2021).

### SARS-CoV-2 Genome Sequencing, Genome Analysis, and Identification of Variants

We sequenced the SARS-CoV-2 genome of >90% of all positive cases in the Houston Methodist healthcare system during the two sampling periods studied. Libraries for whole SARS-CoV-2 genome sequencing were prepared according to version 4 (https://community.artic.network/t/sars-cov-2-version-4-scheme-release/312, last accessed August 19, 2021) of the ARTIC nCoV-2019 sequencing protocol. The semi-automated workflow used has been described previously^2-6^. Sequence reads were generated with an Illumina NovaSeq 6000 instrument.

Viral genomes were assembled with the BV-BRC SARS-Cov2 assembly service (https://www.bv-brc.org/app/ComprehensiveSARS2Analysis, last accessed June 7, 2021, requires registration). The pipeline currently uses seqtk version 1.3-r117 for sequence trimming (https://github.com/lh3/seqtk.git, last accessed December 30, 2021) and minimap version 2.17 for aligning reads against the Wuhan-Hu-1 (NC_045512.2) reference genome. Samtools version 1.11 was used for sequence and file manipulation, where maximum depth and minimum depth parameters in mpileup were set to 8,000 and 3, respectively. iVar version 1.3.1 was used for primer trimming and variant calling. Genetic lineages, VOCs, and variants of interest (VOIs) were identified based on genome sequence data and designated by Pangolin v. 3.1.17 with pangoLEARN module 2021-12-06 (https://cov-lineages.org/resources/pangolin.html, last accessed December 12, 2021). Genome data used in this study have been deposited to GISAID www.gisaid.org.

### S-Gene Target-Failure Assay

An S-gene target-failure (SGTF) assay (TaqPath COVID-19 Combo Kit Thermo Fisher, Inc.), was used as a surrogate marker for the Omicron VOC for some specimens collected between December 18, 2021 and January 5, 2022. From November 1, 2021 onward, only Delta and Omicron were documented to be circulating in metropolitan Houston, based on whole-genome sequence data. Patient samples were first tested in the clinical Molecular Diagnostics Laboratory using a RT-PCR assay with an Emergency Use Authorization as described above. The SARS-CoV-2 positive samples were then tested with the SGTF assay according to the manufacturer’s instructions to infer an Omicron or not-Omicron lineage. That is, the SGTF assay was only performed on samples known to be positive for SARS-CoV-2. Samples yielding amplification of the S-gene were classified as a Delta variant. The SGTF data were validated based on comparing the results with our extensive genome sequence data.

### Patient Metadata and Geospatial Analysis

Patient metadata were acquired from the electronic medical record by standard informatics methods. Figures showing geospatial distribution of spread for Omicron were generated with Tableau version 2021.2.7 (Tableau Software, LLC, Seattle, WA) using patient home address zip codes. A vaccination breakthrough case was defined as a PCR-positive sample from a patient obtained greater than 14 days after full vaccination (e.g., both doses of the Pfizer or Moderna mRNA vaccines) was completed. A booster vaccination breakthrough case was defined as a PCR-positive sample from a patient obtained greater than 14 days after receiving a third vaccine dose. For some cases, manual chart review was conducted to resolve discrepancies or clarify ambiguities.

## Results

### Omicron Epidemiologic Wave

The first Houston Methodist patient infected with an Omicron variant was identified at the end of November 2021, a time when the Delta VOC was responsible for all COVID-19 cases in metropolitan Houston^6^. During this period, the metropolitan area was experiencing a steady decrease in total number of new COVID-19 cases (**Figure 1, Figure 2**).

**Figure 1.**
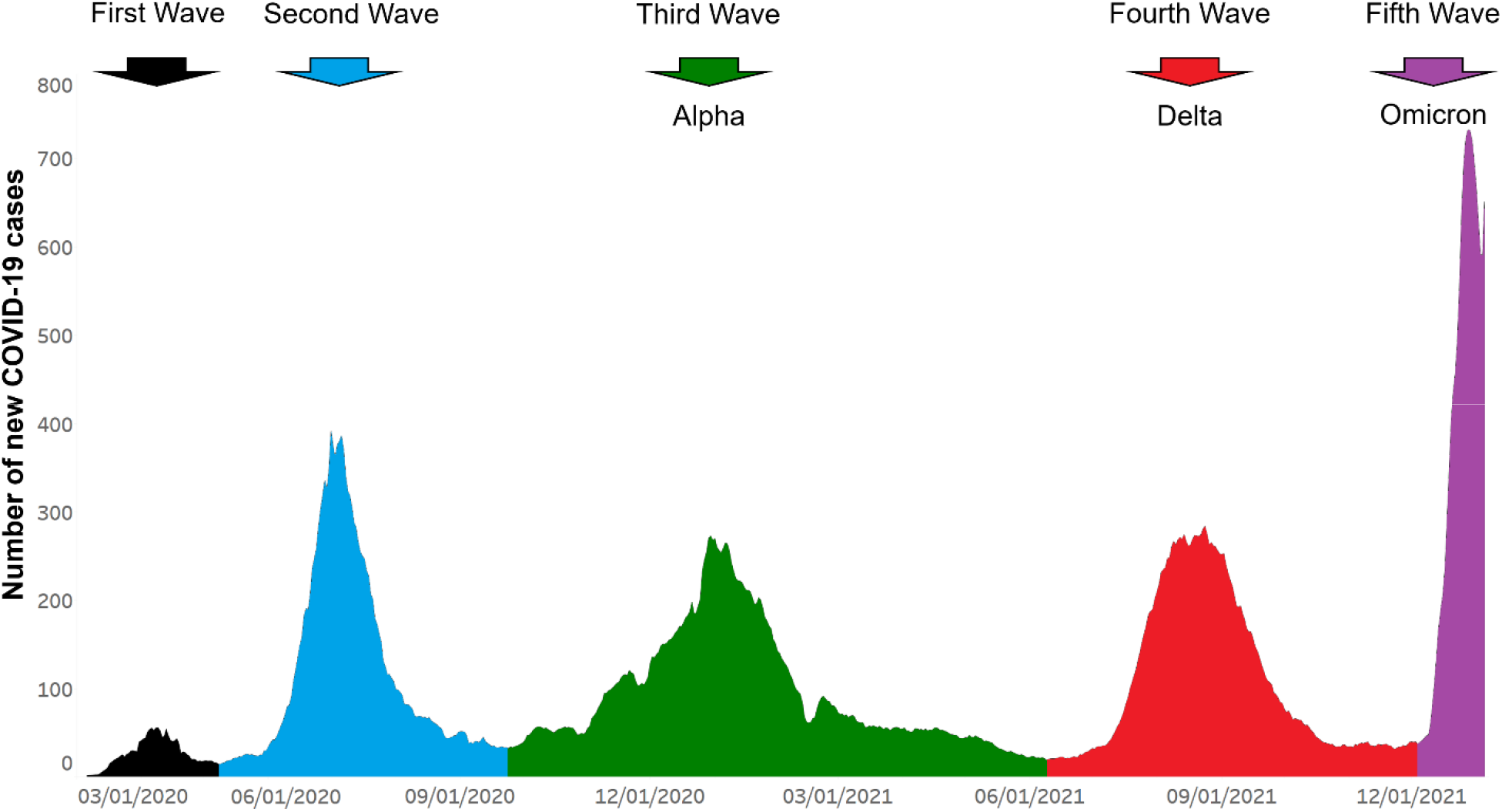
Epidemiologic curve showing five COVID-19 disease waves in Houston Methodist patients. Number of new COVID-19 cases (y-axis) totals are shown as a +/-three-day moving average. Each of the five waves is shown in a different color. The first and second waves were composed of a heterogenous array of SARS-CoV-2 genotypes. The Alpha VOC shown in the third wave, the Delta VOC shown in the fourth, and the Omicron VOC shown in the fifth wave indicate their numeric prominence in those waves. The figure should not be interpreted to mean that all cases in the third, fourth, and fifth waves were caused by Alpha, Delta, and Omicron VOCs, respectively. Rather, they are the dominant single VOCs causing disease in Houston Methodist system patients in those waves. The fifth wave shown includes data through January 5, 2022. The figure was generated with Tableau version 2021.2.7 (Tableau Software, LLC, Seattle, WA), and is a modified version of one presented recently^6^. The curve is essentially superimposable on COVID-19 activity in all metropolitan Houston, Texas.

**Figure 2.**
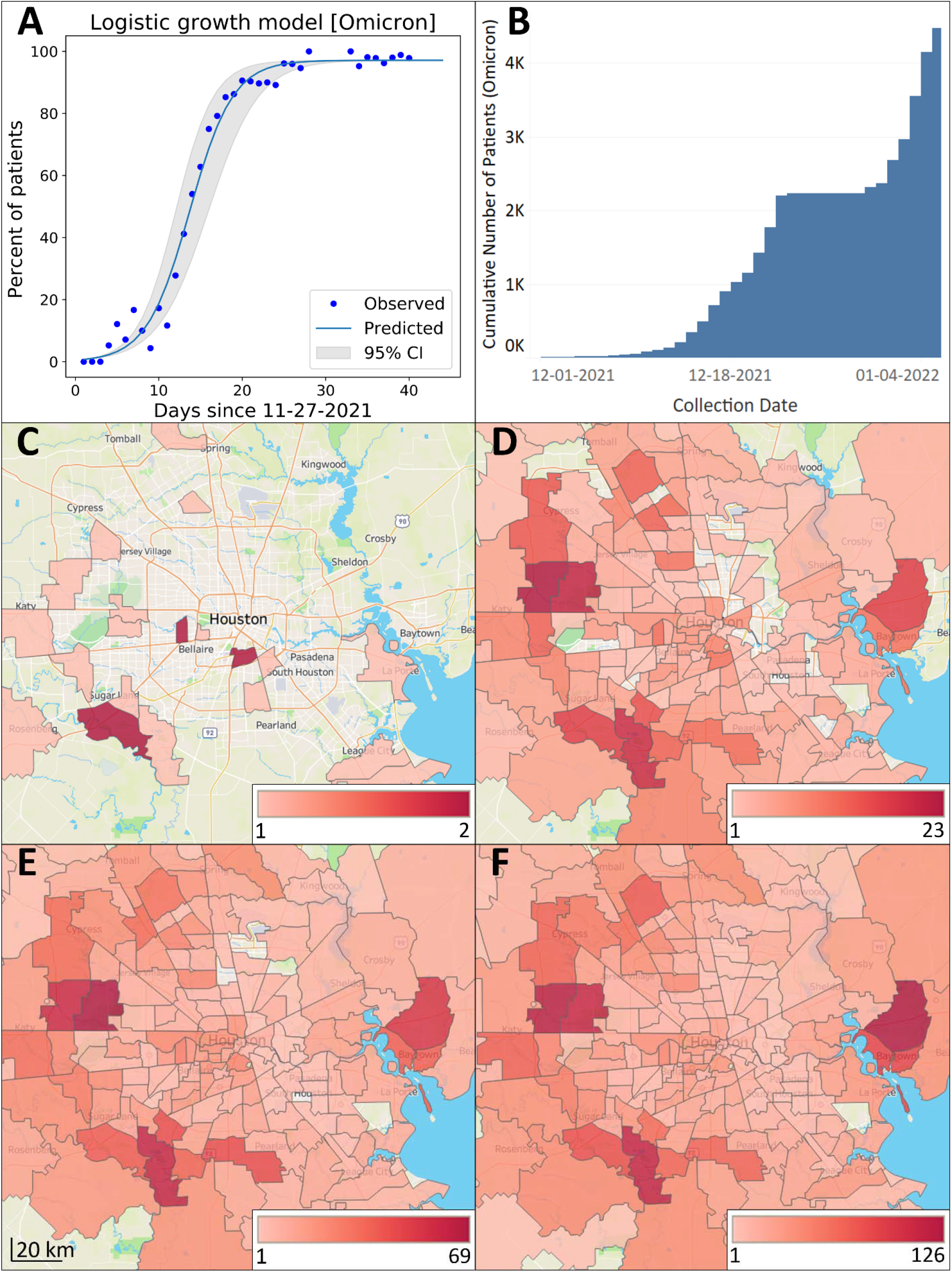
Increase in Omicron frequency over time and distribution in metropolitan Houston. The study time frame was November 27, 2021 through January 5, 2022. **A:** Omicron logistic growth model. The estimated case doubling time is 1.8 days. **B:** Cumulative increase in Omicron during the study period; y-axis is the cumulative number of new COVID-19 Omicron cases. At the end of the study period, Omicron caused 98% of all COVID-19 cases. The plateau between December 24, 2021 and December 30, 2021 exists because we did not sequence samples obtained during this period due to the massive number of daily positive specimens, as described in the Materials and Methods section. **C – F:** Geospatial distribution of Omicron based on home address zip code for each patient. **C**: November 27 – December 6; **D**: November 27 – December 16; **E**: November 27 – December 26; **F**: November 27 – January 5. Note differences in heat map scale for each panel. Figures were generated using Tableau version 2021.2.7. (Tableau Software, LLC, Seattle, WA).

Omicron increased in frequency unusually rapidly over a three-week period in December (**Figure 1, Figure 2)**. By December 23, the genome sequence data showed that Omicron accounted for >90% of all new COVID-19 cases in our healthcare system (**Figure 2**). The estimated case doubling time during this three-week period was approximately 1.8 days (**Figure 2**), which means that Omicron increased in relative frequency approximately three times faster than Delta had increased in our area^6^, an unprecedented trajectory for SARS-COV-2 infections. By January 5, 2022, the Omicron variant caused 98% of all new COVID-19 cases diagnosed in our healthcare system (**Figure 2**). This represents the fifth wave of COVID-19 cases in metropolitan Houston (**Figure 1**).

Consistent with extensive infections caused by Omicron in southern Africa and elsewhere (https://www.cdc.gov/coronavirus/2019-ncov/variants/variant-classifications.html, last accessed December 28, 2021; https://www.gov.uk/government/collections/new-sars-cov-2-variant, last accessed December 28, 2021), several patients had very recent travel histories to countries with a high prevalence of this VOC, suggesting acquisition of virus by some cases from abroad and importation into Houston. However, the vast majority of Omicron patients had no documented travel outside the US and undoubtedly acquired the infection domestically, either in Houston or elsewhere.

To understand the geospatial distribution of Omicron in metropolitan Houston, patient metadata were acquired from the electronic medical record by standard informatics methods, and home address zip codes were used to visualize virus spread (**Figure 2**). The 4,468 Houston Methodist patients infected with Omicron during this period were distributed widely throughout metropolitan Houston, with 259 different zip codes represented (**Figure 2**). The widespread distribution of Omicron in the Houston metroplex in an extremely short period of time reflects the ability of this variant to spread unusually rapidly and effectively between individuals, and cause symptomatic disease.

### Comparison of Omicron, Alpha, and Delta COVID-19 Cases

There is a considerable lack of detailed information about patients with COVID-19 caused by the Omicron VOC, and data are especially lacking for US patients. We compared available metadata for all Houston Methodist patients infected with Omicron, Alpha, and Delta VOCs (**Table 1, Table 2**). The populations differed significantly in many characteristics, including median age, hospital admission rates, maximum respiratory support, rate of vaccine breakthrough, and median length of stay (**Table 1, Table 2**).

**Table 1.**
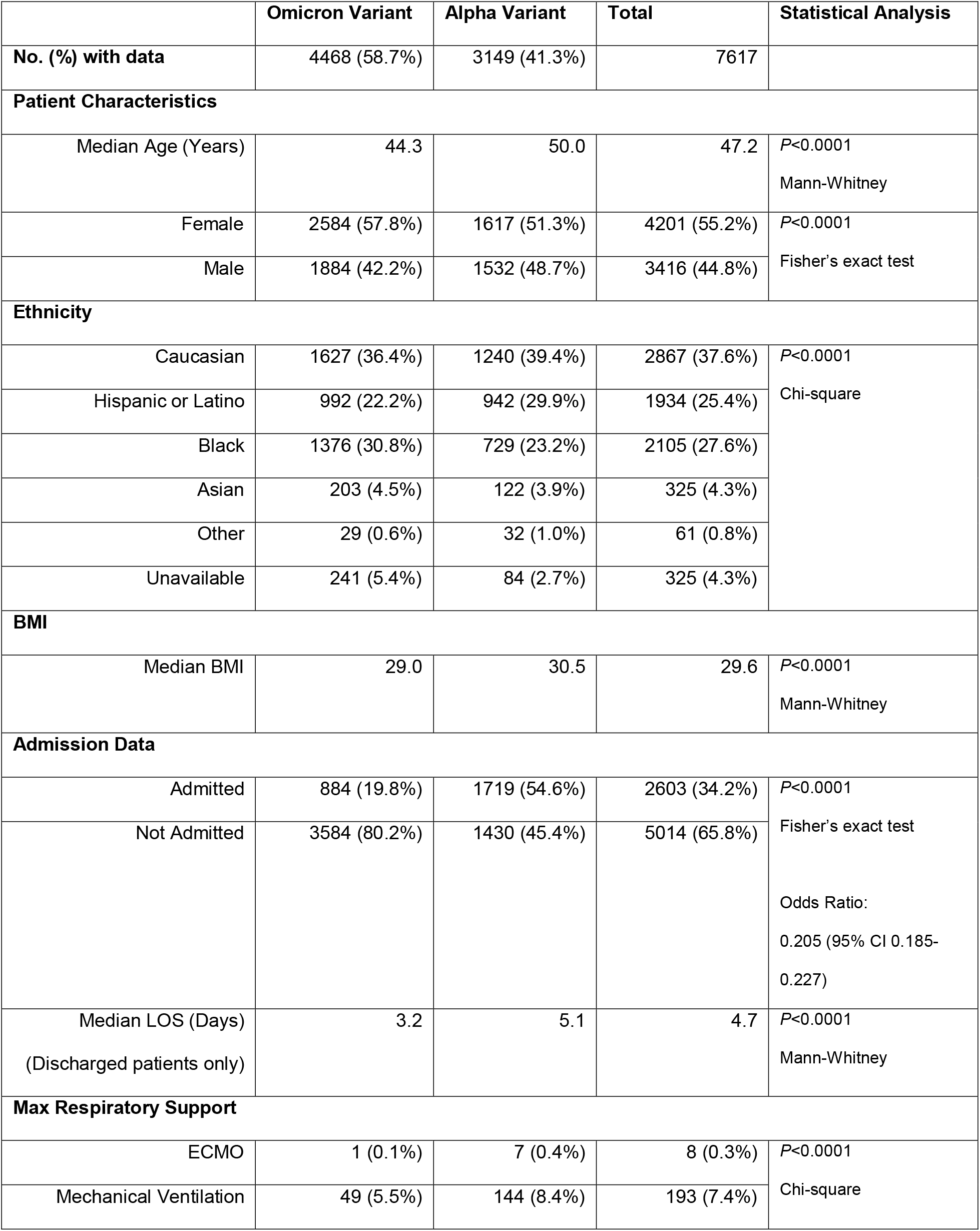
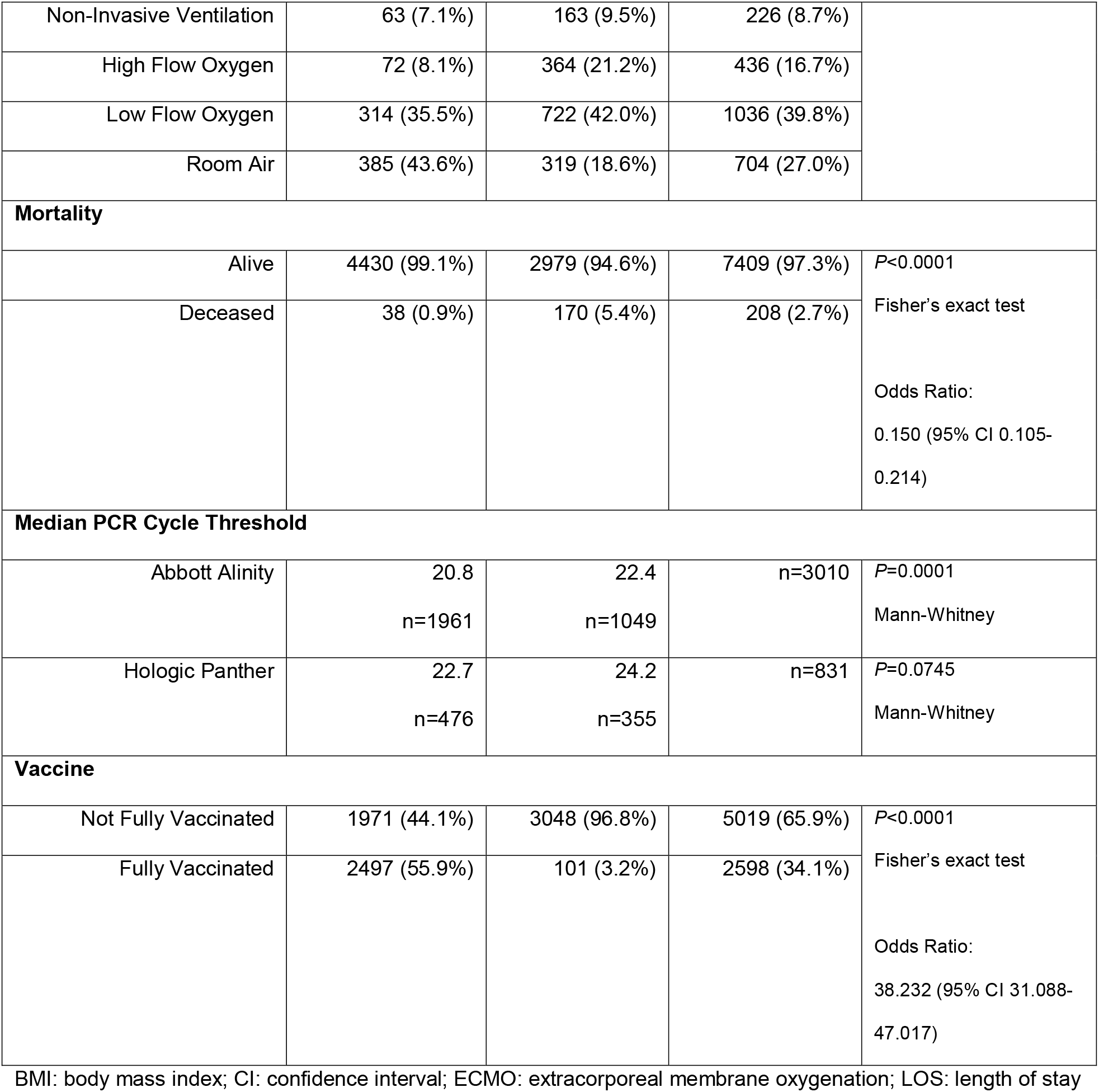
Summary of pertinent patient metadata for 7,617 unique patients infected with Omicron or Alpha variants.

**Table 2.** Summary of pertinent patient metadata for 20,196 unique patients infected with Omicron or Delta variants.

|  | Omicron Variant | Delta Variant | Total | Statistical Analysis |
| --- | --- | --- | --- | --- |
| No. (%) with data | 4468 (22.1%) | 15728 (77.9%) | 20196 |  |
| Patient Characteristics |  |  |  |  |
| Median Age (Years) | 44.3 | 48.3 | 47.6 | P<0.0001<br>Mann-Whitney |
| Female | 2584 (57.8%) | 8123 (51.6%) | 10707 (53.0%) | P<0.0001<br>Fisher's exact test |
| Male | 1884 (42.2%) | 7605 (48.4%) | 9489 (47.0%) |  |
| Ethnicity |  |  |  |  |
| Caucasian | 1627 (36.4%) | 6903 (43.9%) | 8530 (42.2%) | P<0.0001<br>Chi-square |
| Hispanic or Latino | 992 (22.2%) | 4179 (26.6%) | 5171 (25.6%) |  |
| Black | 1376 (30.8%) | 3450 (21.9%) | 4826 (23.9%) |  |
| Asian | 203 (4.5%) | 531 (3.4%) | 734 (3.6%) |  |
| Other | 29 (0.6%) | 112 (0.7%) | 141 (0.7%) |  |
| Unavailable | 241 (5.4%) | 553 (3.5%) | 794 (3.9%) |  |
| BMI |  |  |  |  |
| Median BMI | 29.0 | 29.6 | 29.4 | P<0.0001<br>Mann-Whitney |
| Admission Data |  |  |  |  |
| Admitted | 884 (19.8%) | 6779 (43.1%) | 7663 (37.9%) | P<0.0001<br>Fisher's exact test<br>Odds Ratio:<br>0.326 (95% CI 0.301-0.353) |
| Not Admitted | 3584 (80.2%) | 8949 (56.9%) | 12533 (62.1%) |  |
| Median LOS (Days)<br>(Discharged patients only) | 3.2 | 5.4 | 5.2 | P<0.0001<br>Mann-Whitney |
| Max Respiratory Support |  |  |  |  |
| ECMO | 1 (0.1%) | 19 (0.3%) | 20 (0.3%) | P<0.0001<br>Chi-square |
| Mechanical Ventilation | 49 (5.5%) | 727 (10.7%) | 776 (10.1%) |  |
| Non-Invasive Ventilation | 63 (7.1%) | 641 (9.5%) | 704 (9.2%) |  |
| High Flow Oxygen | 72 (8.1%) | 1796 (26.5%) | 1868 (24.4%) |  |
| Low Flow Oxygen | 314 (35.5%) | 2290 (33.8%) | 2604 (34.0%) |  |
| Room Air | 385 (43.6%) | 1306 (19.3%) | 1691 (22.1%) |  |
| Mortality |  |  |  |  |
| Alive | 4430 (99.1%) | 14889 (94.7%) | 19319 (95.7%) | P<0.0001 |
| Deceased | 38 (0.9%) | 839 (5.3%) | 877 (4.3%) | Fisher's exact test<br>Odds Ratio:<br>0.152 (95% CI 0.110-0.211) |
| Median PCR Cycle Threshold |  |  |  |  |
| Abbott Alinity | 20.8<br>n=1961 | 21.5<br>n=5122 | n=7083 | P<0.0001<br>Mann-Whitney |
| Hologic Panther | 22.7<br>n=476 | 22.6<br>n=1298 | n=1774 | P=0.1606<br>Mann-Whitney |
| Vaccine |  |  |  |  |
| No vaccine | 1815 (40.6%) | 11415 (72.6%) | 13230 (65.5%) | P<0.0001 |
| >7 days past 1st Vaccine | 156 (3.5%) | 494 (3.1%) | 650 (3.2%) | Chi-square |
| >14 days past 2nd Vaccine | 1786 (40.0%) | 3679 (23.4%) | 5465 (27.1%) |  |
| >14 days past 3rd Vaccine | 711 (15.9%) | 140 (0.9%) | 851 (4.2%) |  |

BMI: body mass index; CI: confidence interval; ECMO: extracorporeal membrane oxygenation; LOS: length of stay

Patients infected with Omicron were significantly younger than Alpha and Delta patients (**Table 1, Table 2**). Importantly, Omicron patients were hospitalized significantly less frequently than patients infected with either the Alpha or Delta variants, and had a significantly shorter median hospital length of stay (**Table 1, Table 2**).

We next analyzed Omicron vaccine breakthrough cases (**Table 1, Table 2**). We found 2,497 of the 4,468 total Omicron patients (55.9%) for whom we have whole genome sequence data met the CDC definition of vaccine breakthrough cases (**Table 1, Table 2**). There was no simple relationship between the time elapsed since administration of the second vaccination dose and the date of vaccination breakthrough. These 2,497 patients received either two doses of the Pfizer-BioNTech BNT162b2 (*n* = 1828, 73%) or Moderna mRNA-1273 (*n* = 553, 22%), or one dose of J&J/Janssen JNJ-78436735 (*n* = 115, 5%) vaccine; vaccine type was not specified for one individual. This distribution reflects the majority use of BNT162b2 vaccination doses in our health system. Compared to either Alpha or Delta patients, a significantly greater percentage of patients with breakthrough cases was caused by the Omicron VOC (55.9% compared to 3.2% and 24.3% for Alpha and Delta VOCs, respectively) (**Table 1, Table 2**). We next analyzed individuals with breakthrough cases after receiving a third (booster) dose of either the Pfizer-BioNTech BNT162b2 or Moderna mRNA-1273 vaccine. We found that 711 (15.9%) of the 4,468 Omicron patients met this criteria. Consistent with Omicron causing a significantly increased number of vaccine breakthrough cases, many studies have reported that this variant has reduced sensitivity to antibody neutralization *in vitro*, likely in large part due to the extensive number of amino acid and other structural changes occurring in Omicron spike protein^11-34^.

### Spike-Gene Target-Failure Assay

To estimate Omicron variant frequency in patient samples not yet sequenced, we performed the TaqPath COVID-19 Combo Kit assay (ThermoFisher) on 1,216 samples collected from symptomatic patients between December 18, 2021 and January 5, 2022 In total, 1,093 (90%) of patient samples yielded an RT-PCR result with S-gene target-failure indicative of the Omicron variant. These data are consistent with the increasing frequency of new cases of COVID-19 caused by Omicron in our population (**Figure 2**).

### Discovery of Omicron “Stealth” Sublineage BA.2 in Houston

The Omicron sublineage BA.2 was first identified in November 2021 in Australia in a patient who had traveled to South Africa (https://github.com/cov-lineages/pango-designation/issues/359; last accessed December 30, 2021). This sublineage does not have the full set of polymorphisms characteristic of BA.1 (B.1.1.529) and also has additional mutations unique to it (https://github.com/cov-lineages/pango-designation/issues/361; last accessed December 30, 2021). One important difference is that sublineage BA.2 lacks the spike gene deletion in the region encoding amino acid 69/70 which means that it will not be detected by the SGTF assay. As a consequence, it is sometimes referred to as the Omicron “stealth” variant. We inspected all full genome sequences present in our large database, including specimens obtained from symptomatic patients and asymptomatic individuals, and discovered only two members of the BA.2 sublineage in Houston COVID-19 patients.

## Discussion

This work was conducted to address the relative lack of information about disease character among US patients with COVID-19 caused by the Omicron VOC, and to compare our findings with data available for patients in the Houston Methodist system who had disease caused by the Alpha and Delta VOCs. We describe information relevant to the massive Omicron wave in metropolitan Houston. In three weeks (December 1, 2021 through December 23, 2021), Omicron was first identified in our population and rapidly increased to cause 90% of all new COVID-19 cases, with an unusually fast case doubling time of 1.8 days. Analysis of samples obtained from December 30, 2021 to January 5, 2022 found that at the end of the sampling period Omicron caused 98% of all new COVID-19 cases in our healthcare system.

The study was based on genome sequence analysis of 4,468 Omicron samples taken from socioeconomically, geographically, and ethnically diverse symptomatic patients. Several key findings were made, including (i) the Omicron VOC rapidly increased as a cause of COVID-19 and spread throughout the metroplex in an unusually short period of time, far faster than any other SARS-CoV-2 variant; (ii) Omicron caused significantly more vaccine breakthrough cases than the Alpha or Delta VOCs; (iii) Omicron patients were significantly younger than Alpha or Delta patients; (iv) significantly fewer Omicron patients required hospitalization compared to Alpha and Delta patients; (v) the median length of stay for hospitalized Omicron patients was significantly shorter than for Alpha and Delta patients, and consistent with this observation, on average the maximum respiratory support required for Omicron patients was significantly less than for Alpha or Delta patients. Our findings are largely consistent with many aspects of Omicron data reported from the UK, South Africa, and Canada^8-10, 35-38^, and are consistent with experimental animal infection data suggesting that Omicron causes less severe disease in mice and hamsters^39-43^. This study was facilitated by a comprehensive and integrated population genomics and epidemiology project^2-6^ implemented at the end of February 2020, when the initial COVID-19 case was diagnosed in the Houston Methodist healthcare system.

Several questions arise from our findings, namely the underlying causes for the differences we observe in Omicron compared to Alpha and Delta patients. We believe the data from the extensive studies examining serologic and structural differences in Omicron relative to Alpha and Delta likely contribute to the increased vaccine breakthrough cases observed. It is also possible that waning of immunity is a contributing factor as well. We do not currently have serologic or other data that could address this possibility in our patients. As noted above, ample *in vitro* and animal infection model data have accumulated suggesting that Omicron is less virulent than Delta or Alpha VOC. We speculate that the lower age of Omicron patients may be attributable to a disproportionately greater likelihood of risky behaviors in the younger population, for example less mask wearing and less social distancing. Regardless, additional studies are required to gain more information about factors contributing to the differences between Alpha, Delta, and Omicron patients that we identified in this study.

Because we sequence the genome of approximately 90% of SARS-CoV-2 causing COVID-19 in our diverse Houston Methodist patient population, and have done so for almost two years, we are continuously monitoring the composition of this virus in a major US metroplex. This affords us the opportunity to rapidly assess changes in SARS-CoV-2 population genomic structure in the fourth largest city in the US. However, our study has several limitations. Although we sequenced the genomes of SARS-CoV-2 causing 90% of all Houston Methodist COVID-19 cases in the study period, this sample represents only approximately 5% of cases reported in the metropolitan region. Our patient population will underrepresent some demographic groups, for example homeless individuals and pediatric patients. The samples sequenced in this study were obtained from symptomatic individuals, which means that it is possible that we failed to identify Omicron subvariants or features preferentially represented in asymptomatic individuals. It is likely that our study included some patients where Omicron was detected on hospital admission but was incidental to the primary cause of admission.

The identification of two asymptomatic individuals with the Omicron “stealth” sublineage BA.2 is potentially concerning and stresses the importance of using whole-genome sequencing to study patient samples. This sublineage lacks the spike gene deletion corresponding to amino acids 69 and 70 and is not detected by some commonly used assays. Sublineage BA.2 now accounts for approximately 5% of COVID-19 in the UK, which means that it has the ability to successfully transmit and cause disease^44^. It will be important to determine if this SARS-CoV-2 genotype increases in frequency in metropolitan Houston as additional genome sequencing is conducted on samples from our patient population.

In the aggregate, our data add critical new information to features of Omicron genomic epidemiology and patient characteristics in the US. Further, the present study highlights the importance of analyzing SARS-CoV-2 genome data integrated with patient metadata and stresses the need to continue to do this in near-real time as the Omicron surge continues, the virus evolves, and new variants with potentially altered fitness and biomedically relevant phenotypes are generated. Analyses of this type are also important in the context of vaccine formulation and long COVID, an increasing health and economic problem globally. Finally, the strategy we have used in this and previous studies^2-6^ are readily applicable to future infectious diseases problems that warrant special attention.

## Data Availability

Genome data used in this study have been deposited to GISAID.

https://www.gisaid.org

## Acknowledgments

We thank Drs. Marc Boom and Dirk Sostman for their ongoing support, and Dr. Sasha M. Pejerrey for editorial contributions. The research was supported by the Houston Methodist Academic Institute Infectious Diseases Fund and many generous Houston philanthropists. The funders had no role in the design and conduct of the study; collection, management, analysis, and interpretation of the data; preparation, review, or approval of the manuscript; and decision to submit the manuscript for publication.

We declare that we have no conflict of interest.

## Author Contributions

P.A.C., R.J.O., S.W.L., and J.M.M. had full access to all study data and take responsibility for the integrity of the data and the accuracy of the data analysis; concept and design by J.M.M., P.A.C., R.J.O., and S.W.L; data acquisition, analysis, or interpretation by all authors; drafting of the manuscript by all authors; statistical analysis by P.A.C.; funding obtained by J.M.M. and J.J.D.; and overall supervision by J.M.M. P.A.C., R.J.O., and S.W.L. contributed equally and are co-first authors.

## Notes

Funding: This project was supported by the Houston Methodist Academic Institute Infectious Diseases Fund; and in part with funds from the National Institute of Allergy and Infectious Diseases, National Institutes of Health, Department of Health and Human Services, under Contract No. 75N93019C00076 (J.J.D.).

### Competing Interest Statement

The authors have declared no competing interest.

### Funding Statement

This project was supported by the Houston Methodist Academic Institute Infectious Diseases Fund; and supported in whole or in part with federal funds from the National Institute of Allergy and Infectious Diseases, National Institutes of Health, Department of Health and Human Services, under Contract No. 75N93019C00076 (J.J.D. and R.O.).

### Author Declarations

This work was approved by the Houston Methodist Research institutional review board (IRB1010-0199).

### Summary of Updates

Tables and figures updated with additional data.

## References

[1] Dhar MS, Marwal R, Vs R, Ponnusamy K, Jolly B, Bhoyar RC, et al.: Genomic characterization and epidemiology of an emerging SARS-CoV-2 variant in Delhi, India.Science 2021, 374:995–999

[2] Long SW, Olsen RJ, Christensen PA, Bernard DW, Davis JJ, Shukla M, Nguyen M, Saavedra MO, Yerramilli P, Pruitt L, Subedi S, Kuo HC, Hendrickson H, Eskandari G, Nguyen HAT, Long JH, Kumaraswami M, Goike J, Boutz D, Gollihar J, McLellan JS, Chou CW, Javanmardi K, Finkelstein IJ, Musser JM: Molecular Architecture of Early Dissemination and Massive Second Wave of the SARS-CoV-2 Virus in a Major Metropolitan Area. mBio 2020, 11

[3] Musser JM, Olsen RJ, Christensen PA, Long SW, Subedi S, Davis JJ, Gollihar J: Rapid, widespread, and preferential increase of SARS-CoV-2 B.1.1.7 variant in Houston, TX, revealed by 8,857 genome sequences. medRxiv 2021 [Preprint]. doi:2021.2003.2016.21253753

[4] Olsen RJ, Christensen PA, Long SW, Subedi S, Hodjat P, Olson R, Nguyen M, Davis JJ, Yerramilli P, Saavedra MO, Pruitt L, Reppond K, Shyer MN, Cambric J, Gadd R, Thakur RM, Batajoo A, Finkelstein IJ, Gollihar J, Musser JM: Trajectory of Growth of Severe Acute Respiratory (SARS-CoV-2) Syndrome Coronavirus 2 Variants in Houston, Texas, January through May 2021, Based on 12,476 Genome Sequences. Am J Pathol 2021, Oct;191(10):1754–1773

[5] Long SW, Olsen RJ, Christensen PA, Subedi S, Olson R, Davis JJ, Saavedra MO, Yerramilli P, Pruitt L, Reppond K, Shyer MN, Cambric J, Finkelstein IJ, Gollihar J, Musser JM: Sequence Analysis of 20,453 SARS-CoV-2 Genomes from the Houston Metropolitan Area Identifies the Emergence and Widespread Distribution of Multiple Isolates of All Major Variants of Concern. Am J Pathol 2021, Nov 11;S0002-9440(21)00480-6

[6] Christensen PA, Olsen RJ, Long SW, Subedi S, Davis JJ, Hodjat P, Walley DR, Kinskey JC, Saavedra MO, Pruitt L, Reppond K, Shyer MN, Cambric J, Gadd R, Thakur RM, Batajoo A, Mangham R, Pena S, Trinh T, Yerramilli P, Nguyen M, Olson R, Snehal R, Gollihar J, Musser JM: Delta Variants of SARS-CoV-2 Cause Significantly Increased Vaccine Breakthrough COVID-19 Cases in Houston, Texas. Am J Pathol 2021 Nov 11:S0002-9440(21)00480-6

[7] Christensen PA, Olsen RJ, Long SW, Snehal R, Davis JJ, Saavedra MO, Reppond K, Shyer MN, Cambric J, Gadd R, Thakur RM, Batajoo A, Mangham R, Pena S, Trinh T, Kinskey JC, Williams G, Olson R, Gollihar J, Musser JM: Early signals of significantly increased vaccine breakthrough, decreased hospitalization rates, and less severe disease in patients with COVID-19 caused by the Omicron variant of SARS-CoV-2 in Houston, Texas. medRxiv 2022. [Preprint]. doi:2021.2012.2030.21268560

[8] Viana R, Moyo S, Amoako DG, Tegally H, Scheepers C, Lessells RJ, et al.: Rapid epidemic expansion of the SARS-CoV-2 Omicron variant in southern Africa. medRxiv 2021. [Preprint]. doi:2021.2012.2019.21268028

[9] Elliott P, Bodinier B, Eales O, Wang H, Haw D, Elliott J, Whitaker M, Jonnerby J, Tang D, Walters CE, Atchison C, Diggle PJ, Page AJ, Trotter AJ, Ashby D, Barclay W, Taylor G, Ward H, Darzi A, Cooke GS, Chadeau-Hyam M, Donnelly CA: Rapid increase in Omicron infections in England during December 2021: REACT-1 study. medRxiv 2021. [Preprint]. doi:2021.2012.2022.21268252

[10] Sheikh Ak, Steven; Woolhouse, Mark; McMenamin, Jim; Robertson, Chris. : Severity of Omicron variant of concern and vaccine effectiveness against symptomatic disease: national cohort with nested test negative design study in Scotland. The University of Edinburgh 2021

[11] Meng B, Ferreira I, Abdullahi A, Kemp SA, Goonawardane N, Papa G, Fatihi S, Charles O, Collier D, Collaboration C-NBC-, Consortium TGtPJ, Choi J, Hyeon Lee J, Mlcochova P, James L, Doffinger R, Thukral L, Sato K, Gupta RK: SARS-CoV-2 Omicron spike mediated immune escape, infectivity and cell-cell fusion. bioRxiv 2021. [Preprint]. doi:2021.2012.2017.473248

[12] Zeng C, Evans JP, Qu P, Faraone J, Zheng Y-M, Carlin C, Bednash JS, Zhou T, Lozanski G, Mallampalli R, Saif LJ, Oltz EM, Mohler P, Xu K, Gumina RJ, Liu S-L: Neutralization and Stability of SARS-CoV-2 Omicron Variant. bioRxiv 2021. [Preprint]. doi:2021.2012.2016.472934

[13] Jacobsen H, Strengert M, Maass H, Ynga Durand MA, Kessel B, Harries M, Rand U, Abassi L, Kim Y, Lueddecke T, Hernandez P, Ortmann J, Heise J-K, Castell S, Gornyk D, Gloeckner S, Melhorn V, Lange B, Dulovic A, Haering J, Junker D, Schneiderhan-Marra N, Poehlmann S, Hoffmann M, Krause G, Cicin-Sain L: Diminished neutralization responses towards SARS-CoV-2 Omicron VoC after mRNA or vector-based COVID-19 vaccinations. medRxiv 2021. [Preprint]. doi:2021.2012.2021.21267898

[14] Eggink D, Andeweg SP, Vennema H, van Maarseveen N, Vermaas K, Vlaemynck B, Schepers R, van Gageldonk-Lafeber AB, van den Hof S, Reusken Cbem, Knol MJ: Increased risk of infection with SARS-CoV-2 Omicron compared to Delta in vaccinated and previously infected individuals, the Netherlands, 22 November to 19 December 2021. medRxiv 2021. [Preprint]. doi:2021.2012.2020.21268121

[15] Edara V-V, Manning KE, Ellis M, Lai L, Moore KM, Foster SL, Floyd K, Davis-Gardner ME, Mantus G, Nyhoff LE, Bechnack S, Alaaeddine G, Naji A, Samaha H, Lee M, Bristow L, Hussaini L, Ciric CR, Nguyen P-V, Gagne M, Roberts-Torres J, Henry AR, Godbole S, Grakoui A, Sexton M, Piantadosi A, Waggoner JJ, Douek DC, Anderson EJ, Rouphael N, Wrammert J, Suthar MS: mRNA-1273 and BNT162b2 mRNA vaccines have reduced neutralizing activity against the SARS-CoV-2 Omicron variant. bioRxiv 2021. [Preprint]. doi:2021.2012.2020.473557

[16] Zou j, Xia H, Xie X, Kurhade C, Machado RR, Weaver SC, Ren P, Shi P-Y: Neutralization against Omicron SARS-CoV-2 from previous non-Omicron infection. bioRxiv 2021. [Preprint]. doi:2021.2012.2020.473584

[17] Ikemura N, Hoshino A, Higuchi Y, Taminishi S, Inaba T, Matoba S: SARS-CoV-2 Omicron variant escapes neutralization by vaccinated and convalescent sera and therapeutic monoclonal antibodies. medRxiv 2021. [Preprint]. doi:2021.2012.2013.21267761

[18] Dejnirattisai W, Shaw RH, Supasa P, Liu C, Stuart AS, Pollard AJ, Liu X, Lambe T, Crook D, Stuart DI, Mongkolsapaya J, Nguyen-Van-Tam JS, Snape MD, Screaton GR: Reduced neutralisation of SARS-CoV-2 omicron B.1.1.529 variant by post-immunisation serum. Lancet 2021. (in press) doi:10.1016/s0140-6736(21)02844-0

[19] Cameroni E, Saliba C, Bowen JE, Rosen LE, Culap K, Pinto D, et al.: Broadly neutralizing antibodies overcome SARS-CoV-2 Omicron antigenic shift. Nature Research Briefing 2021. [Preprint]. doi:2021.2012.2012.472269

[20] Liu L, Iketani S, Guo Y, Chan JF-W, Wang M, Liu L, Luo Y, Chu H, Huang Y, Nair MS, Yu J, Chik KK-H, Yuen TT-T, Yoon C, To KK-W, Chen H, Yin MT, Sobieszczyk ME, Huang Y, Wang HH, Sheng Z, Yuen K-Y, Ho DD: Striking Antibody Evasion Manifested by the Omicron Variant of SARS-CoV-2. Nature Research Briefing 2021. [Preprint]. doi:2021.2012.2014.472719

[21] Planas D, Saunders N, Maes P, Benhassine FG, Planchais C, Porrot F, Staropoli I, Lemoine F, Pere H, Veyer D, Puech J, Rodary J, Bolland WH, Buchrieser J, Baele G, Dellicour S, Raymenants J, Gorissen S, Geenen C, Vanmechelen B, Wawina T, Marti J, Cuypers L, Seve A, Hocqueloux L, Prazuck T, Loriere ES, Rey F, Bruel T, Mouquet H, Andre E, Schwartz O: Considerable escape of SARS-CoV-2 variant Omicron to antibody neutralization. Nature Research Briefing 2021. [Preprint]. doi:2021.2012.2014.472630

[22] Andrews N, Stowe J, Kirsebom F, Toffa S, Rickeard T, Gallagher E, Gower C, Kall M, Groves N, O’Connell A-M, Simons D, Blomquist PB, Zaidi A, Nash S, Aziz NIBA, Thelwall S, Dabrera G, Myers R, Amirthalingam G, Gharbia S, Barrett JC, Elson R, Ladhani SN, Ferguson N, Zambon M, Campbell CN, Brown K, Hopkins S, Chand M, Ramsay M, Bernal JL: Effectiveness of COVID-19 vaccines against the Omicron (B.1.1.529) variant of concern. medRxiv 2021. [Preprint]. doi:2021.2012.2014.21267615

[23] Yu X, Wei D, Xu W, Li Y, Li X, Zhang X, Qu J, Yang Z, Chen E: Reduced sensitivity of SARS-CoV-2 Omicron variant to booster-enhanced neutralization. medRxiv 2021. [Preprint]. doi:2021.2012.2017.21267961

[24] Cele S, Jackson L, Khan K, Khoury DS, Moyo-Gwete T, Tegally H, Scheepers C, Amoako D, Karim F, Bernstein M, Lustig G, Archary D, Smith M, Ganga Y, Jule Z, Reedoy K, Cromer D, San JE, Hwa S-H, Giandhari J, Blackburn JM, Gosnell BI, Karim SSA, Hanekom W, NGS-SA, Team C-K, von Gottberg A, Bhiman J, Lessells RJ, Moosa M-YS, Davenport MP, de Oliveira T, Moore PL, Sigal A: SARS-CoV-2 Omicron has extensive but incomplete escape of Pfizer BNT162b2 elicited neutralization and requires ACE2 for infection. medRxiv 2021. [Preprint]. doi:2021.2012.2008.21267417

[25] Cao Y, Wang J, Jian F, Xiao T, Song W, Yisimayi A, Huang W, Li Q, Wang P, An R, Wang J, Wang Y, Niu X, Yang S, Liang H, Sun H, Li T, Yu Y, Cui Q, Liu S, Yang X, Du S, Zhang Z, Hao X, Shao F, Jin R, Wang X, Xiao J, Wang Y, Xie XS: B.1.1.529 escapes the majority of SARS-CoV-2 neutralizing antibodies of diverse epitopes. Nature Research Briefing 2021. [Preprint]. doi:2021.2012.2007.470392

[26] Hansen CH, Schelde AB, Moustsen-Helms IR, Emborg H-D, Krause TG, Moelbak K, Valentiner-Branth P, Institut TIDPGaSS: Vaccine effectiveness against SARS-CoV-2 infection with the Omicron or Delta variants following a two-dose or booster BNT162b2 or mRNA-1273 vaccination series: A Danish cohort study. medRxiv 2021. [Preprint]. doi:2021.2012.2020.21267966

[27] Syed AM, Ciling A, Khalid MM, Sreekumar B, Kumar GR, Silva I, Milbes B, Kojima N, Hess V, Shacreaw M, Lopez L, Brobeck M, Turner F, Spraggon L, Taha TY, Tabata T, Chen IP, Ott M, Doudna JA: Omicron mutations enhance infectivity and reduce antibody neutralization of SARS-CoV-2 virus-like particles. medRxiv 2021. [Preprint]. doi:2021.2012.2020.21268048

[28] Sheward DJ, Kim C, Ehling RA, Pankow A, Castro Dopico X, Martin DP, Reddy ST, Dillner J, Karlsson Hedestam GB, Albert J, Murrell B: Variable loss of antibody potency against SARS-CoV-2 B.1.1.529 (Omicron). bioRxiv 2021. [Preprint]. doi:2021.2012.2019.473354

[29] Haveri A, Solastie A, Ekström N, Österlund P, Nohynek H, Nieminen T, Palmu AA, Melin M: Neutralizing antibodies to SARS-CoV-2 Omicron variant after 3rd mRNA vaccination in health care workers and elderly subjects and response to a single dose in previously infected adults. medRxiv 2021. [Preprint]. doi:2021.2012.2022.21268273

[30] Arien KK, Heyndrickx L, Michiels J, Vereecken K, Van Lent K, Coppens S, Pannus P, Martens GA, Van Esbroeck M, Goossens ME, Marchant A, Bartholomeeusen K, Desombere I: Three doses of the BNT162b2 vaccine confer neutralising antibody capacity against the SARS-CoV-2 B.1.1.529 (Omicron) variant of concern. medRxiv 2021. [Preprint]. doi:2021.2012.2023.21268316

[31] Willett BJ, Grove J, MacLean O, Wilkie C, Logan N, De Lorenzo G, et al.: The hyper-transmissible SARS-CoV-2 Omicron variant exhibits significant antigenic change, vaccine escape and a switch in cell entry mechanism. medRxiv 2022. [Preprint]. doi:2022.2001.2003.21268111

[32] Boschi C, Colson P, Bancod A, Moal V, La Scola B: Omicron variant escapes therapeutic mAbs contrary to eight prior main VOC. bioRxiv 2022. [Preprint]. doi:2022.2001.2003.474769

[33] Dejnirattisai W, Huo J, Zhou D, Zahradník J, Supasa P, Liu C, et al.: Omicron-B.1.1.529 leads to widespread escape from neutralizing antibody responses. bioRxiv 2021. [Preprint]. doi:10.1101/2021.12.03.471045

[34] Banerjee A, Lew J, Kroeker A, Baid K, Aftanas P, Nirmalarajah K, Maguire F, Kozak R, McDonald R, Lang A, Gerdts V, Straus SE, Gilbert L, Li AX, Mozafarihasjin M, Walmsley S, Gingras A-C, Wrana JL, Mazzulli T, Colwill K, McGeer AJ, Mubareka S, Falzarano D: Immunogenicity of convalescent and vaccinated sera against clinical isolates of ancestral SARS-CoV-2, beta, delta, and omicron variants. bioRxiv 2022. [Preprint]. doi:2022.2001.2013.475409

[35] Ulloa AC, Buchan SA, Daneman N, Brown KA: Early estimates of SARS-CoV-2 Omicron variant severity based on a matched cohort study, Ontario, Canada. medRxiv 2021. [Preprint]. doi:2021.2012.2024.21268382

[36] Wolter N, Jassat W, Walaza S, Welch R, Moultrie H, Groome M, Amoako DG, Everatt J, Bhiman JN, Scheepers C, Tebeila N, Chiwandire N, du Plessis M, Govender N, Ismail A, Glass A, Mlisana K, Stevens W, Treurnicht FK, Makatini Z, Hsiao N-y, Parboosing R, Wadula J, Hussey H, Davies M-A, Boulle A, von Gottberg A, Cohen C: Early assessment of the clinical severity of the SARS-CoV-2 Omicron variant in South Africa. medRxiv 2021. [Preprint]. doi:2021.2012.2021.21268116

[37] SARS-CoV-2 variants of concern and variants under investigation in England Technical briefing: Update on hospitalisation and vaccine effectiveness for Omicron VOC-21NOV-01 (B.1.1.529) UK Health Security: UK Health Security, 2021.

[38] Davies M-A, Kassanjee R, Rousseau P, Morden E, Johnson L, Solomon W, et al.: Outcomes of laboratory-confirmed SARS-CoV-2 infection in the Omicron-driven fourth wave compared with previous waves in the Western Cape Province, South Africa. medRxiv 2022. [Preprint]. doi:2022.2001.2012.22269148

[39] Bentley EG, Kirby A, Sharma P, Kipar A, Mega DF, Bramwell C, Penrice-Randal R, Prince T, Brown JC, Zhou J, Screaton GR, Barclay WS, Owen A, Hiscox JA, Stewart JP: SARS-CoV-2 Omicron-B.1.1.529 Variant leads to less severe disease than Pango B and Delta variants strains in a mouse model of severe COVID-19. bioRxiv 2021. [Preprint]. doi:2021.2012.2026.474085

[40] Abdelnabi R, Foo CS, Zhang X, Lemmens V, Maes P, Slechten B, Raymenants J, André E, Weynand B, Dallemier K, Neyts J: The omicron (B.1.1.529) SARS-CoV-2 variant of concern does not readily infect Syrian hamsters. bioRxiv 2021. [Preprint]. doi:2021.2012.2024.474086

[41] Diamond M, Peter H, Tadashi M, Kiyoko I-H, Shun I, Maki K, et al.: The SARS-CoV-2 B.1.1.529 Omicron virus causes attenuated infection and disease in mice and hamsters Nature Portfolio 2022. 29 December 2021. [Preprint]. doi:https://doi.org/10.21203/rs.3.rs-1211792/v1

[42] McMahan K, Giffin V, Tostanoski L, Chung B, Siamatu M, Suthar M, Halfmann P, Kawaoka Y, Piedra-Mora C, Martinot A, Kar S, Andersen H, Lewis MG, Barouch DH: Reduced Pathogenicity of the SARS-CoV-2 Omicron Variant in Hamsters. bioRxiv 2022. [Preprint]. doi:2022.2001.2002.474743

[43] Yuan S, Ye Z-W, Liang R, Tang K, Zhang AJ, Lu G, Ong CP, Poon VK-M, Chan CC-S, Mok BWY, Qin Z, Xie Y, Sun H, Tsang JO-L, Yuen TT-T, Chik KK-H, Chan CC-Y, Cai J-P, Luo C, Lu L, Yip CC-Y, Chu H, To KK-W, Chen H, Jin D-Y, Yuen K-Y, Chan JFW: The SARS-CoV-2 Omicron (B.1.1.529) variant exhibits altered pathogenicity, transmissibility, and fitness in the golden Syrian hamster model. bioRxiv 2022. [Preprint]. doi:2022.2001.2012.476031

[44] SARS-CoV-2 variants of concern and variants under investigation in England Technical briefing 34. UK Health Security Agency. January 14, 2022.

